# The Iberian Roma genetic variant server; population structure, susceptibility to disease and adaptive traits

**DOI:** 10.1101/2023.08.25.23294490

**Authors:** Fabiola Mavillard, Javier Pérez-Florido, Francisco M Ortuño, Amador Valladares, Miren L Álvarez-Villegas, Gema Roldán, Rosario Carmona, Manuel Soriano, Santiago Susarte, Pilar Fuentes, Daniel López-López, Ana María Nuñez-Negrillo, Alejandra Carvajal, Yolanda Morgado, Daniel Arteaga, Rosa Ufano, Pablo Mir, Juan F Gamella, Joaquín Dopazo, Carmen Paradas, Macarena Cabrera-Serrano

**Affiliations:** Instituto de Biomedicina de Sevilla (IBiS), Hospital Universitario Virgen del Rocío/CSIC/Universidad de Sevilla. Sevilla, Spain; Centro Investigación Biomédica en Red Enfermedades Neurodegenerativas (CIBERNED). Instituto de salud Carlos III. Sevilla, Spain; Plataforma Andaluza de Medicina Computacional, Fundación Progreso y Salud (FPS), Hospital Virgen del Rocío, Sevilla, Spain; Grupo de medicina computacional de sistemas, Instituto de Biomedicina de Sevilla (IBiS), Hospital Virgen del Rocío, Sevilla, Spain; Nodo de Genómica Funcional, (INB-ELIXIR-es), Fundación Progreso y Salud (FPS), Hospital Virgen del Rocío, Sevilla 41013, Spain; Bioinformática en Enfermedades raras (BiER), Centro de Investigación Biomédica en Red de Enfermedades Raras (CIBERER), Instituto de salud Carlos III. Sevilla, Spain; Departamento de Ingeniería de Computadores, Automática y Robótica, Universidad de Granada, Granada, Spain; Servicio de Urgencias de Atención Primaria. Distrito Sevilla. Sevilla, Spain; Centro de Servicios Sociales, Negociado de Servicios Especializados. Ayuntamiento de Sevilla, Sevilla, Spain; Departamento de Enfermería, Facultad de Ciencias de la Salud, Universidad de Granada, Granada, Spain; Departamento de Neurología, Hospital Virgen de las Nieves, Granada, Spain; Departamento de Neurología, Hospital Virgen de Valme, Sevilla, Spain; Centro de Salud Utrera Sur, Sevilla; Centro de Salud Poligono Sur, Sevilla; Unidad de Trastornos del Movimiento, Servicio de Neurología y Neurofisiología Clínica. Hospital Universitario Virgen del Rocío, Sevilla, Spain; Departamento de Medicina, Facultad de Medicina, Universidad de Sevilla, Seville, Spain; Departamento de Antropología Social, Universidad de Granada, Spain; Unidad Enfermedades Neuromusculares, Servicio de Neurología y Neurofisiología Clínica. Hospital Universitario Virgen del Rocío, Sevilla, Spain

**Author notes:** **Correspondance to:** Macarena Cabrera-Serrano, Department of Neurology. Hospital Universitario Virgen del Rocio. 41013 Sevilla. Spain, Carmen Paradas, Department of Neurology. Hospital Universitario Virgen del Rocio. 41013 Sevilla. Spain, Joaquin Dopazo, Grupo de medicina computacional de sistemas, Instituto de Biomedicina de Sevilla (IBiS), Hospital Virgen del Rocío,41013 Sevilla, Spain. These authors have equally contributed to this work (Fabiola Mavillard, Javier Pérez-Florido).

## Abstract

The Roma are the most numerous ethnic minority in Europe. The Iberian Roma arrived in the Iberian Peninsula five centuries ago and still today, they keep a strong group identity. Demographic and cultural reasons lie behind a high rate of Mendelian disease often related to founder variants. We have analysed exome data from 119 Iberian Roma individuals collected from 2018 to 2020. A database of variant frequency has been implemented (IRPVS) and made available online. We have analysed the carrier rate of founder private alleles as well as pathogenic variants present in the general population. Significant enrichment in structural variants involving gene clusters related to keratinization and epidermal growth suggest that evolutive mechanisms have developed towards climate and environmental adaptation. IRPVS can be accessed at http://irpvs.clinbioinfosspa.es/

**AUTHOR SUMMARY:** Reference data is necessary for the correct interpretation of genetic studies. Although most genetic variants are present in all populations, ancestry has an important impact in the genetic background. For that reason databases of genetic variant in populations are developed specifically for different ethnicities, being an important tool for genetic diagnosis. The Roma are the most numerous ethnic minority in Europe. In this study we have collected samples from healthy Roma individuals from Iberian descent and implemented a database of genetic variant to facilitate genetic diagnosis in this population. Analysis of structural variants that are specific to the Iberian Roma not found in other healthy population for which genetic data are available suggest evolution towards environmental adaptation.

## INTRODUCTION

In recent years, we have witnessed a thorough transformation of the knowledge and understanding of genetic disease. With massive parallel sequencing becoming available and its cost decreasing rapidly, the number of known disease genes has exponentially grown. So has our understanding of their effects, leading to progressive improvement of methods and protocols for assessing and classifying the variants according to their disease-causing potential. The presence of a large amount of rare variants in populations has become evident[1–3]. Since most pathogenic variants are rare in the general population, and variant frequency being fundamental information during the variant-assessment process, implementation of public repositories of variant frequency, such as Gnomad[4], has become an indispensable tool for genetic diagnosis [5]. Accumulating evidence shows disease susceptibility is associated with low-frequency variants[6, 7] that are often specific to populations of distinct ancestries [3, 7–11], which makes necessary having population-specific reference data. There are many examples of disease-causing variants that are specific to a population and may explain a large proportion of the cases of a disease in this group[12–14]. There is also the opposite situation in which a benign variant, frequent in a population due to founder effects, is rare in another population. If data from the wrong control population group is consulted, the assumed low frequency could be taken in support of the variant as a potential candidate for causing disease[15]. Using data from an incorrect reference group could not only prevent a correct diagnosis, but lead to misdiagnosis [16]. Commonly used protocols for variant-assessing highlight the necessity of using reference frequency data from the specific population, due to the presence of benign variants common in some populations and very rare in others [17]. The absence of normative data from some minorities causes patients to receive more uncertain results from genetic studies than patients from other populations groups well represented in public repositories [18]. This is particularly important for founder populations and isolated communities with a high number of private variants[19]. Numerous calls, from scientific and non-scientific sources, have been made to include ethnic minorities data in genomic studies for the sake of knowledge as well as equality of opportunities to access tailored medicine [16, 20, 21] (https://www.genome.gov/news/news-release/Genomic-databases-weakened-by-lack-of-non-European-populations),(https://theconversation.com/how-the-genomics-health-revolution-is-failing-ethnic-minorities-86385), (https://blogs.scientificamerican.com/voices/we-need-more-diversity-in-our-genomic-databases/) and (https://www.genome.gov/news/news-release/nih-awards-38-million-dollars-to-improve-utility-of-polygenic-risk-scores-in-diverse-populations). Some population specific databases have been developed[3, 22–28] and large databases of genetic variants are often subdivided by ethnic background or geographical origin[4]. Yet, for some populations there no reference data publicly available to be consulted. Expanded carrier screening programs, designed to include the whole population of a country or region, is less effective to detect pathogenic variants that are private to a minority for which there is not normative data [29]. For the Roma, being the largest ethnic minority in Europe, there is no available information of genetic variant frequency, which renders them in a situation of disadvantage and inequality.

According to estimates of the European Commission, the European Roma, also known as Romani, or by their own ethnonyms, Roma, Sinti, Kale, etc., comprise 10 to 12 million people (https://commission.europa.eu/strategy-and-policy/policies/justice-and-fundamental-rights/combatting-discrimination/roma-eu/roma-equality-inclusion-and-participation-eu_en). Although there is no official census, the population size has gone through rapid expansion over the last decades[30]. From a genetic point of view, the Roma is a population with high incidence of genetic disease and private features that make it not comparable to the general European population, similarly to other founder populations, like Ashkenazi or Finnish. The Roma descend from a reduced number of ancestors[31]. The original population founders; referred to as proto-Roma, were located to north India by linguistic and later genetic studies. Migrations through Persia, and the Balkans on their way to Europe can be traced through genetic admixture[30, 32]. A high number of founder events together with endogamy, are some of the reasons behind the high rate of Mendelian disease in the Roma. Carrier rates for some diseases have been estimated to be as high as 5 to 20%[33]. Ancient founder mutations are present in all Roma groups, despite being geographically diverse[34]. Younger mutations are restricted to specific groups, like the *CTDP1* IVS6+389C→T causing congenital cataracts facial dysmorphism neuropathy to a few Balkan Roma groups[35] and the *BIN1* p.Arg234Cys variant, causing centronuclear myopathy, to the Iberian Roma, with a carrier rate of 3.5%[36].

The Iberian Roma are one of the most numerous Roma communities in Europe. It is estimated that the Roma first arrived in the Iberian peninsula around 1425 AD and had higher levels of admixture with the host population than other settlements[30]. Still, there is a strong cultural, historical and familial background that preserves a well-defined group identity to the present day.

The knowledge of the genetic background of the Roma through a systematic study of healthy volunteers will improve the diagnostic rate for diseases and thus treatment, prevention of complications, genetic counselling and prevention of further cases in a family. Besides, the identification of disease-causing mutations with high carrier rate among the population, along with an effective information program would enable informed decision-making of the affected individuals.

Prompted by the difficulties in variant assessing and genetic diagnosis in the Roma invidividuals, in this study we have collected blood samples from healthy volunteers of Roma ethnicity for exome sequencing and built a database of genetic variants in the Iberian Roma population that has been made available online. We have performed a comprehensive analysis of variants, autozygosity and evolutionary traces.

## RESULTS

One hundred and nineteen DNA samples were included in the study. Of those, 88 were newly recruited for the study and rest were archive samples. The average age was 42 years (range 18 to 80) and 73% were women. Geographical origin was diverse including nine different Autonomous Communities in Spain and Portugal. Province of origin of the individual and both parents was available for 76 samples, with an intergenerational mobility rate of 51%.

A mean coverage of 105.84x±19.30x was obtained for the whole experiment with the majority of target bases (mean of 95.19%±3.18%) covered at ≥30x. These results ensured the quality of the variant calling process.

### 1. Iberian Roma population variant server (IRPVS) database

IRPVS is an open resource available at http://irpvs.clinbioinfosspa.es. The set of counts of high-quality variants identified in the Iberian Roma cohort have been uploaded into it for search purposes and will be updated with data from new samples as they become available.

#### The IRPVS interface

The initial screen (Figure 1a) requires the acceptance of the “Terms and conditions for the use of the IRPVS database” (http://irpvs.clinbioinfosspa.es/downloads/IRPVSTermsAndConditions_use.pdf) before any operation is run.

**Figure 1:**
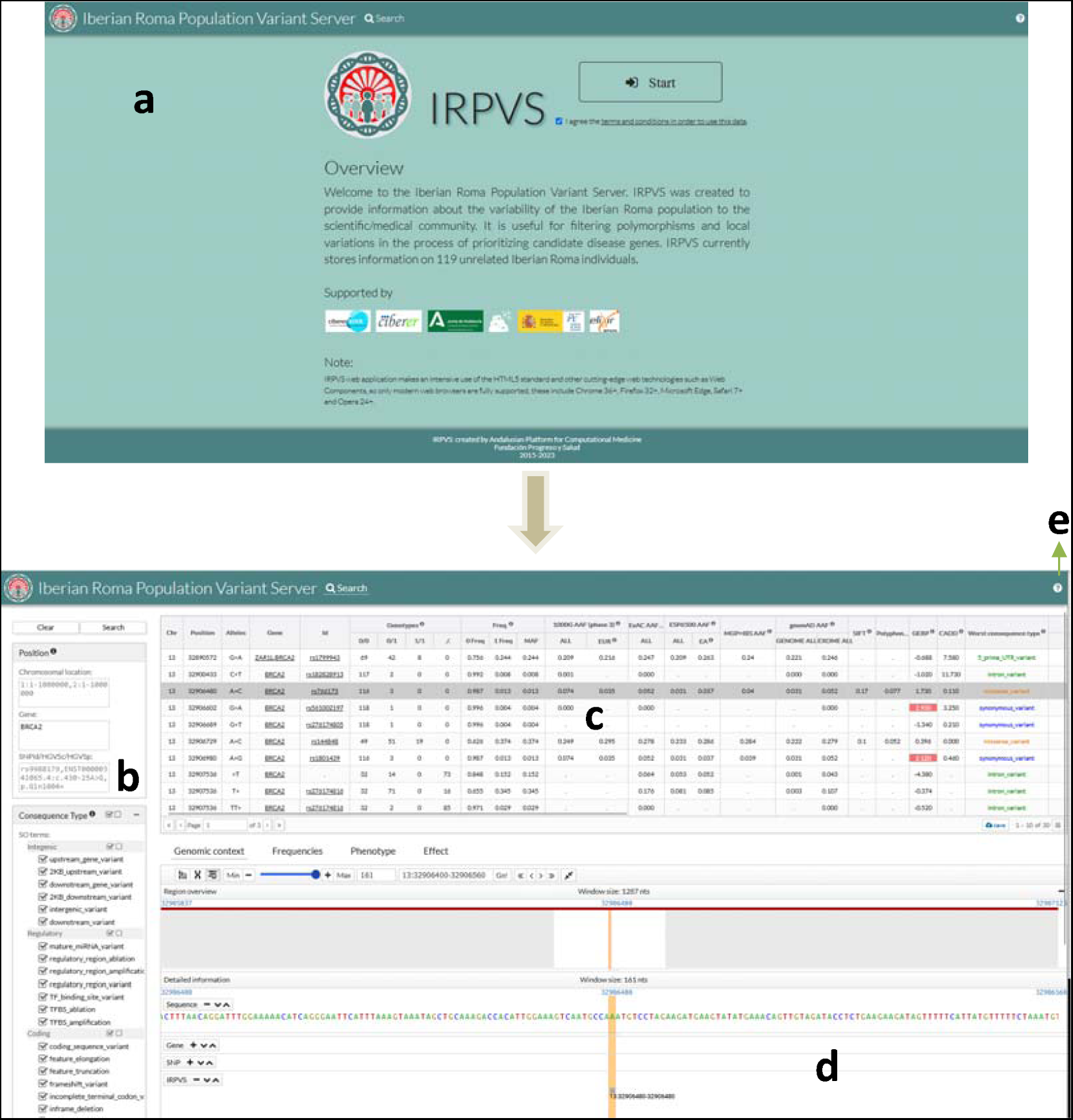
The Iberian Roma Population Variant Server (IRPGS) database. **a** Initial IRPVS page. **b** Query panel in the Search option. **c** List of variants found in the Iberian Roma population within the selected region along with complementary information on impact, conservation, other’s population frequencies and phenotype. **d** Genomic browser that displays the selected variant in its genomic context. e Help icon with links to Full documentation, source code, database version and contact details.

The search option allows querying the IRPVS database. In the left panel (Figure 1b) queries can be done by gene symbol, chromosomal regions, dbSNP entry, HGVS transcript nomenclature (HGVSc) or the HGVS protein nomenclature (HGVSp). The search can be filtered by Sequence Ontology terms for the variation consequences. Also, variants can be highlighted using different types of scores such as SIFT[37], Polyphen[38], CADD [39] or GERP[40].

The results of the query (Figure 1c) include a list of the positions for which variation has been found in the IRPVS along with complementary data such as: chromosome, position, reference allele and alternative allele, genotype and allelic frequencies in the IRPVS database and impact and conservation indexes (e.g. SIFT, Polyphen, CADD, Gerp) . Allelic frequency of each variant in other population databases including 1000 genomes project, ESP (“Exome Variant Server · Bio.Tools”), gnomAD v3.1.2 and the Spanish population from the MGP database (CSVS[28]), is also shown. The most deleterious consequence type found and annotation of the canonical transcript among all transcripts for a given variant are also shown. For variants present in the ClinVar[41] or COSMIC[42], the associated phenotype is included. ClinVar and COSMIC are annotated interactively on each query using the Cellbase[43] webservices. Also a visualization of the variant in the genomic context of the selected variant is shown (Figure 1d) in a parallel window based on the Genome Maps browser[44]. Additionally, some extra detailed information can be found on the population frequencies observed for the variant, the phenotype or the effect. Full documentation of IRPVS and a link to source code can be found at http://irpvs.clinbioinfosspa.es (Figure 1e).

### 2. The Relationship of the Iberian Roma with other populations

The comparison of the genetic variants present in the Iberian Roma (cohort of study) with all other subpopulations present in 1000 Genomes (including general Spanish population- IBS, among others) and MGP (Spanish Medical Genome Project cohort), shows proximity of the Iberian Roma to European and South Asian populations, in good agreement with previous genetic and demographic studies[30, 45]. The principal component analysis (PCA) shows the Iberian Roma cluster being well delimited and in close continuity with the general Spanish cluster and European populations (Figure 2). When compared to specific non-Iberian Roma cluster[46] and taking into account the migrant group, a very close proximity and an overlapping can be observed with samples from Eastern Europe and North/Western Europe respectively (Supplementary Figure 1).

**Figure 2.**
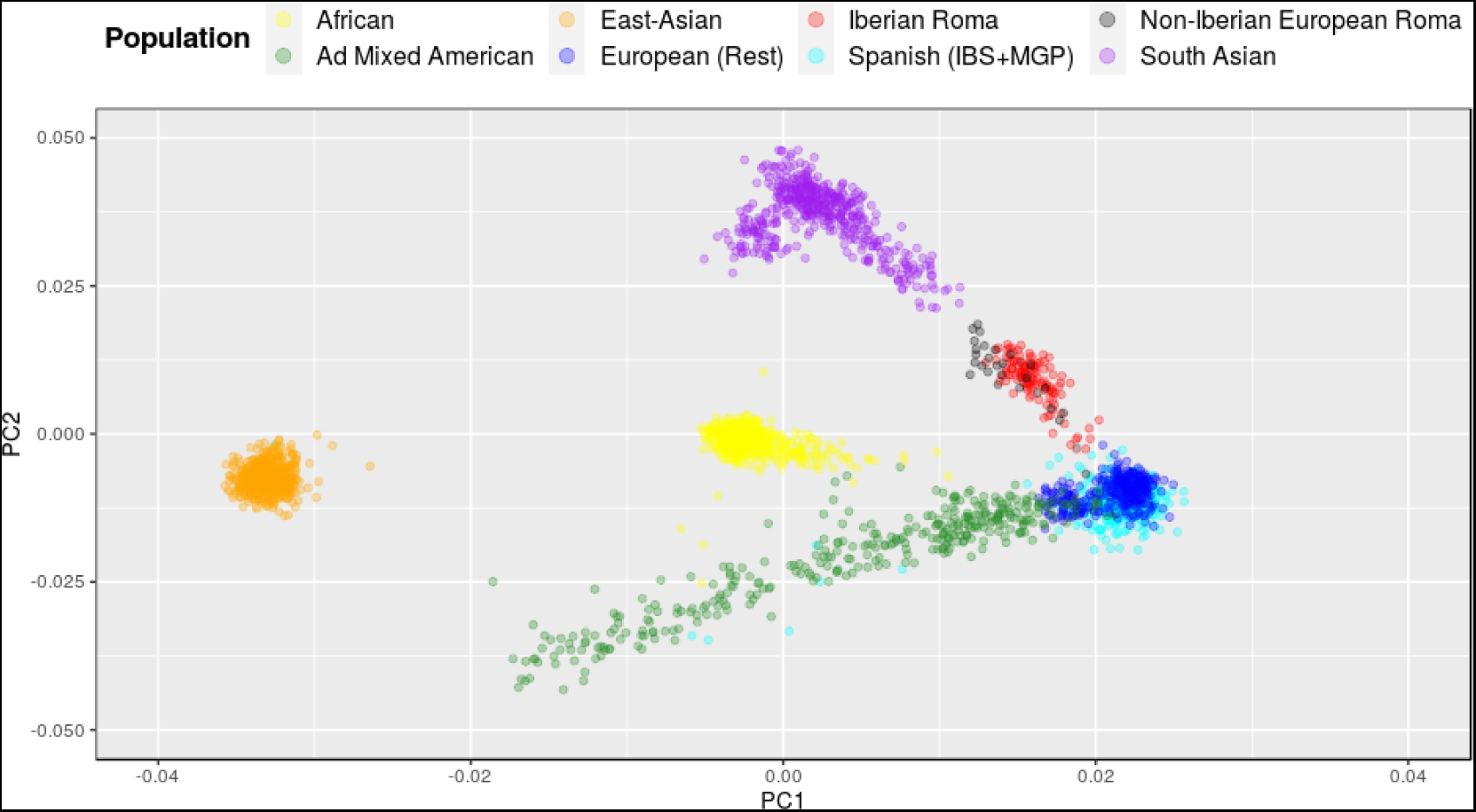
Principal Component analysis (PCA). PCA analysis performed using the 119 Iberian Roma samples included in this study as well as MGP, Thousand Genomes subpopulations and non-iberian European Roma restricted to common captured regions in Iberian Roma and MGP populations and exonic regions from RefSeq.

### 3. Variant analysis

#### Variability distribution in the Iberian Roma population

Total numbers of variants observed in the cohort of study are shown in Table 1. Of note, 39.5% of the SNVs identified are homozygous (38% when considering both SNVs and Indels) compared to 36.6% of homozygous SNVs previously described for the Spanish MGP population[3]. Table 2 shows the number of variants identified in the Iberian Roma in this study comparing it to Spanish MGP, 1000 genomes (including IBS) and other non-iberian Roma[46] populations. For comparative purposes, we have calculated average numbers per individual in each population with similar results amongst populations in the case of detected variants. When taking into account singleton variants, the Iberian Roma population shows the lowest average value, being in line with previous studies that have shown less singletons in Iberian Roma when compared to other populations[47].

**Table 1.** Variants observed in the Iberian Roma population. Total number of variants and average per individual in the Iberian Roma population restricted to the union of the two exome captures. A variant is labelled as singleton if it is present only in a single individual in the Iberian Roma cohort

|  | TOTAL<br>VARIANTS | AVERAGE<br>VARIANTS<br>PER<br>INDIVIDUAL | AVERAGE<br>VARIANTS PER<br>INDIVIDUAL<br>(HOMOZYGOUS) |
| --- | --- | --- | --- |
| Positions with SNVs and Indels | 179,597 | 40,740.83 | 15,564.47 |
| Monoallelic positions | 174,799 | 40,547.46 | 15,564.47 |
| Multiallelic positions | 4,798 | 193.37 | 0 |
| SNVs | 163,497 | 36,647.5 | 14,490.66 |
| Indels | 25,280 | 4,286.7 | 1,073.82 |
| Singletons | 47,063 | 395.49 | 14.87 |

**Table 2.** Total and average numbers of variants in different populations. Total number of variants and average per individual in the Iberian Roma compared to other populations, restricted to common captured regions in Iberian Roma and MGP populations and exonic regions from RefSeq. To be fair in the comparison of singleton variants and their average value amongst cohorts, the whole set of 2,920 individuals have been taken into account, that is, a variant is labelled as a singleton if it is present in a single individual but not in any other of the remaining 2,919 individuals.

|  | POPULATION |  |  |  |  |
| --- | --- | --- | --- | --- | --- |
|  | MGP (N=267) | MGP+IBS (N=374) | MGP+1000g (N=2,771) | IB Roma (N=119) | Non-IB Roma (N=30) |
| SNVs | 165,140 | 21,850 | 1,037,545 | 97,098 | 71,233 |
| Indels | - | 2,035* | 6,665* | 5,671 | 3,238 |
| Singletons | 37,962 | 55,734 | 520,779 | 9,402 | 4,540 |
| Average SNVs per individual | 18,077.93 | 19,256.82 | 23,102.69 | 21,698.74 | 22,624.53 |
| Average Indels per individual | - | 531.17* | 552.06* | 835.00 | 820.76 |
| Average singletons per individual | 142.18 | 149.02 | 187.94 | 79.01 | 151.33 |

\* For MGP population, indels were not reported. Therefore these samples were not taken into account for indel metrics (total and average per individual)

#### Private variants

Variants that are private to the Iberian Roma were obtained by subtracting those present in Spanish MGP and 1000 genomes (Table 3). Approximately, 13% of the private variants (SNVs and Indels) are homozygous (9% homozygous SNVs and 17% homozygous indels) compared to 7% of the Spanish private MGP population where only SNVs are reported [3]. We found that 56% of the private variants of the IRPVS (including SNP and Indels) are present in only one individual (singletons), much lower than the 85% found in the Spanish MGP that includes only SNPs[3]. Moreover, 1% of the IRPVS population shares 45% of the private variants in the cohort, whereas for the Spanish MGP 1% of the population shares 5% of the private variants. These differences are higher when we look at all the variants in the population, including private and non-private. To illustrate this notion, Figure 3 compares Iberian Roma and Spanish MGP populations in terms of variants shared by growing fractions of the corresponding population showing that in the Iberian Roma, private variants are shared by more individuals compared to Spanish MGP.

**Figure 3.**
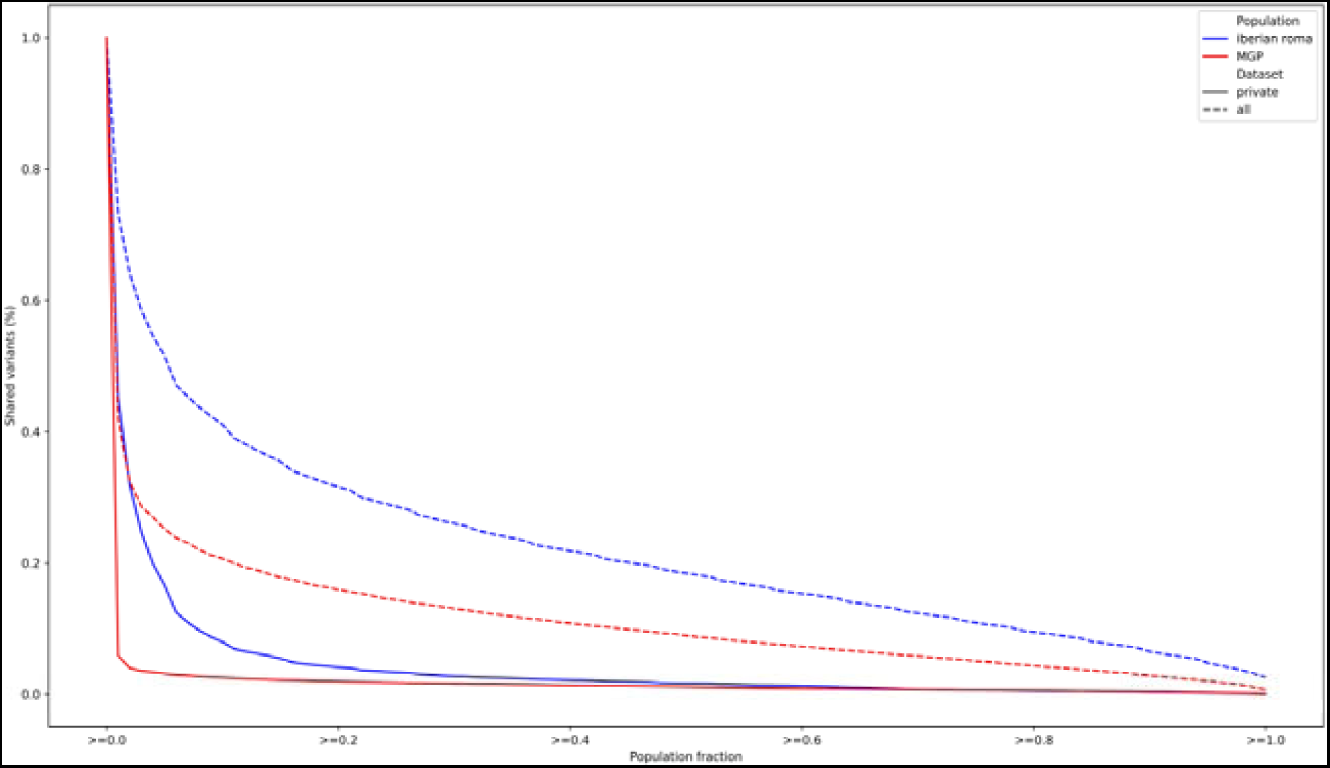
Variants shared by growing fraction of Iberian Roma and MGP populations. Two comparisons are shown: (i) private Iberian Roma variants obtained by substracting those present in MGP or 1000 genomes (blue line) and private MGP variants obtained by substracting those present in 1000 genomes (red line) and (ii) Iberian Roma and MGP variants (blue and red dashed lines, respectively). In both cases, the analysis was restricted to common captured regions in Iberian Roma and MGP populations and exonic regions from RefSeq.

**Table 3.** Private variants. Variants observed in the Iberian Roma not present in the general Spanish population (MGP) and 1000 genomes. A variant is labelled as singleton if it is present only in a single individual in the private variant dataset.

|  | PRIVATE<br>VARIANTS | AVERAGE<br>PRIVATE PER<br>INDIVIDUAL | AVERAGE<br>PRIVATE<br>VARIANTS PER<br>INDIVIDUAL<br>(HOMOZYGOUS) |
| --- | --- | --- | --- |
| Positions with SNVs and Indels | 18,434 | 733.22 | 94.26 |
| Monoallelic positions | 17,975 | 721.08 | 94.26 |
| Multiallelic positions | 456 | 12.14 | 0 |
| SNVs | 15,095 | 388.07 | 35.13 |
| Indels | 4,054 | 357.29 | 59.13 |
| Singletons | 10,744 | 90.29 | 2.52 |

In a similar fashion, Figure 4 illustrates the increment of new variants as more individuals are included in our cohort, showing saturation of the curve is not reached. Non-private variants grow more rapidly while private variants contribute to the growth to a lesser extent. When we look specifically at private variants, we can see how the contribution of singletons is similar to the contribution of polymorphic (present in more than one individual) private variants, showing parallel lines of growth. This is strikingly different from what was described for the Spanish MGP population, where the private variants grow mainly from addition of singletons with a small contribution of polymorphic variants [3].

**Figure 4.**
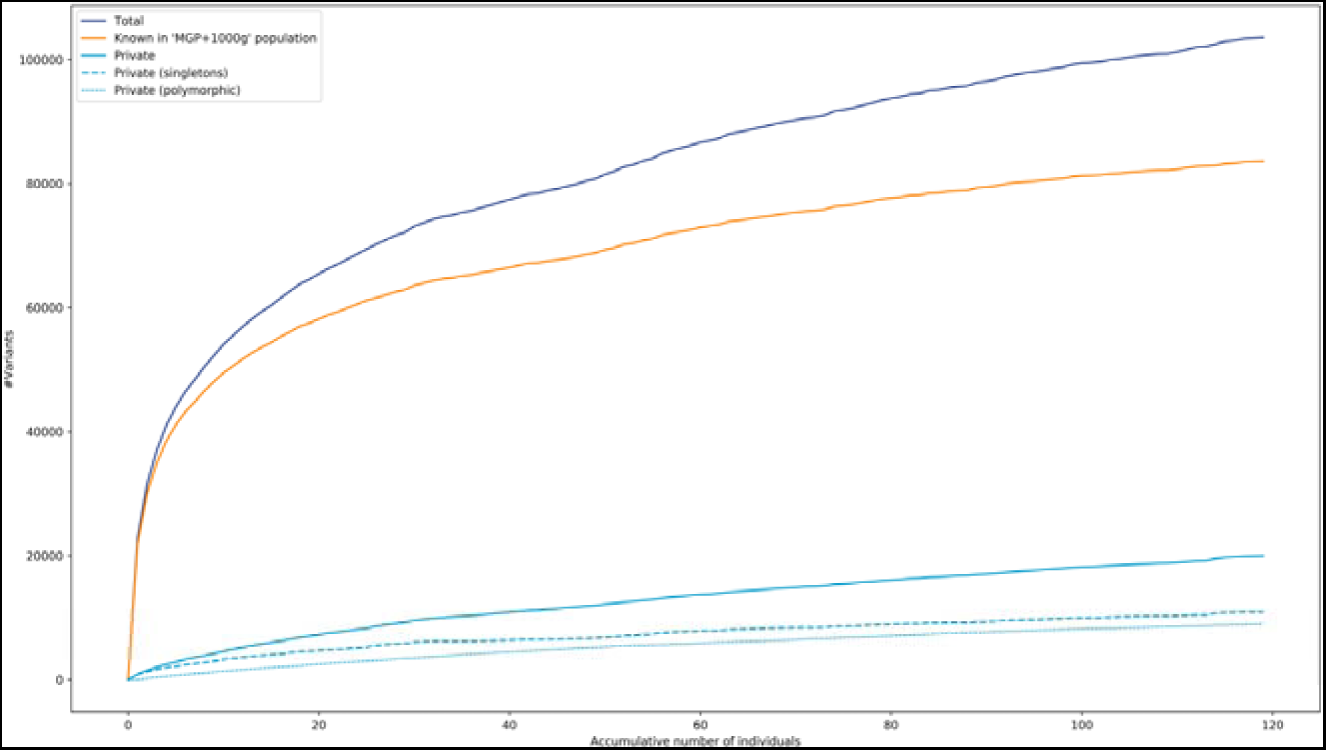
Accumulative number of variants contributed by individuals. Accumulative number of variants contributed by individuals. As the number of individuals in the population grows, so does the total number of variants (dark blue line). Variants that are not private to the Iberian Roma account for a major part of this growth (orange line). Private variants show a slower pattern of growth contributing to a lesser extent to the total (light blue line). Private variants are decomposed in singletons, present in a single individual (light blue dashed line) and polymorphic private variants, present in more than one individual (light blue dotted line)

To check, in terms of Roma population, the uniqueness of private Iberian Roma variants, the intersection of such dataset with non-Iberian Roma population was assessed, arising a significant set of 12,863 SNPs and 3,189 Indels that are unique of Iberian Roma population (84% of the private Iberian Roma variants). The distribution of variants in IRPVS according to the main consequence type categories given by *Cellbase* shows a lower proportion of non-disruptive variants that might change protein effectiveness (MODERATE category) and a greater proportion of non-coding variants (MODIFIER category) when compared to other populations (MGP+1000G) in the same regions of interest (Figure 5). Interestingly, when considering only private Roma variants (i.e. not present in MGP+1000G population), there is a low proportion of non-disruptive and mostly harmless variants (MODERATE and LOW categories, respectively). On the contrary, there is a high proportion of non-coding variants (MODIFIER category) and sites with high (disruptive) impact in the protein (HIGH category), which includes splice-sites, stop-loss and frameshift variants (see Supplementary Figure 2 for details on each category). These results are also in line with previous studies that have shown more proportion of deleterious variants in Roma compare to non-Roma populations[47].

**Figure 5.**
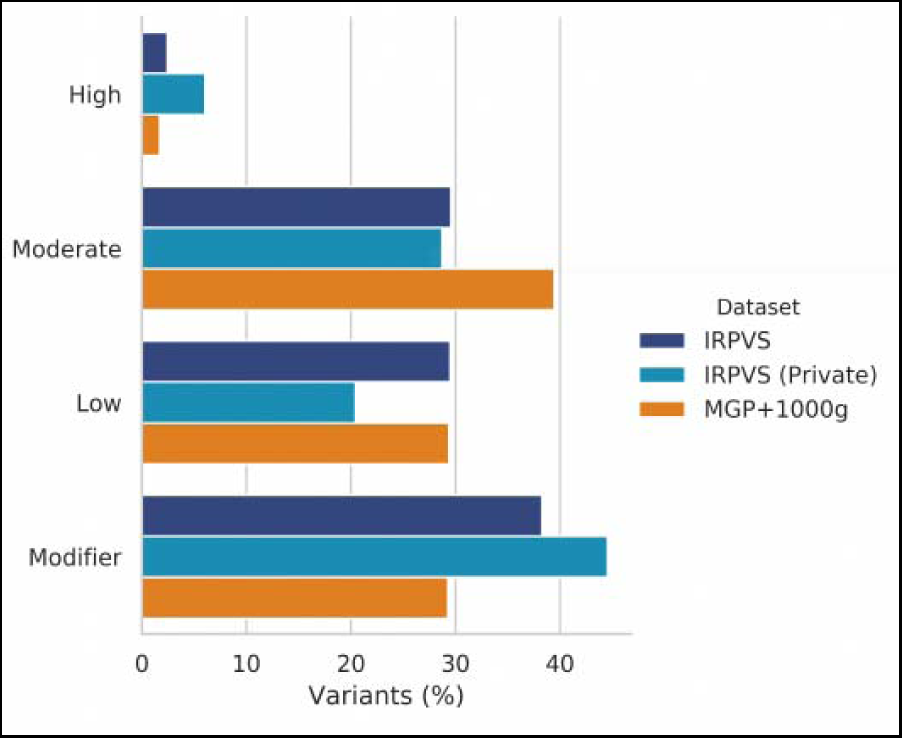
Distribution of variants per consequence type. Distribution of variants in IRPVS (dark blue bar) and MGP+1000G (orange bar) according to Ensemble’s worst consequence type (the worst effect that the variant has on the set of transcripts) obtained through *Cellbase* and restricted to the union of the two exome captures of IRPVS data. Iberian Roma variants not present in MGP+1000G population (light blue bar) is also shown. Consequence type is classified in one of the four main categories (HIGH, MODERATE, LOW and MODIFER) which reflects the severity or impact of the variant consequence.

### 4. Autozygosity

Runs of homozygosity are more common and on average larger in the Iberian Roma than other populations. Specifically, the Iberian Roma population had an average of 14.3 RoH (>1Mb) per sample whereas this metric decreases to 10.8 RoH for the 1000G and MGP. RoH were also significantly longer (*P < 2.2e^-16^*, Wilcoxon non-parametric test) for the Iberian Roma, with an average length of 8.3Mb compared to 3.78Mb for 1000G and MGP populations combined (Figure 6a). When we analyse the length of RoH per chromosome we see an even distribution involving all autosomes similarly (Figure 6b).

**Figure 6.**
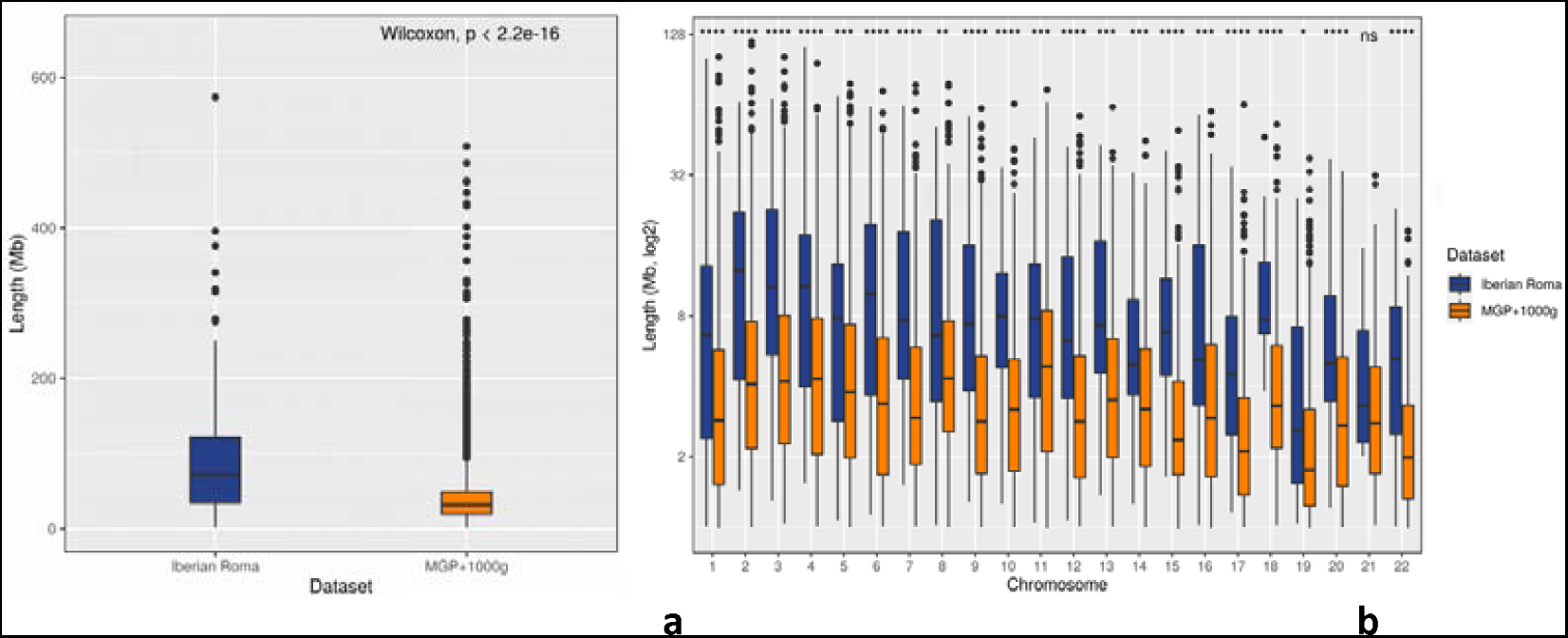
Runs of Homozygosity (RoH). **a** Average length of RoH per sample in the IRPVS compared to a reference population (MGP + 1000G). **b** Length of RoH per chromosome in the IRPVS compared to MGP + 1000G. The following convention for symbols indicating statistical significance were used: (i) ns: p > 0.05, (ii) *: p <= 0.05, (iii) **: p <= 0.01, (iv) *** p <= 0.001 and (v) **** p <= 0.0001

Another interesting metric is the proportion of the autosomal genome in RoH over the specified target regions, termed as F_ROH_. The F_ROH_ provides an accurate estimation of the total inbreeding coefficient of each individual[48]. The Iberian Roma population revealed a mean F_ROH_ of 4.78% whereas only 1.6% was reached by the reference population (1000 Genomes and MGP datasets).

### 5. Deviation from Hardy-Weinberg (HW) equilibrium

A total of 95,765 SNVs from sites with a single alternative allele in the Iberian Roma cohort restricted to common captured regions in Iberian Roma and Spanish MGP populations and exonic regions from Refseq[49] were tested for possible deviations from the HW equilibrium. As a result, 5,818 (6.1%, Supplementary Table 1) variants deviated significantly from the equilibrium (P < 0.05), 1,468 (1.5%) variants could not be computed for numerical restrictions (NA) and the rest of variants (88,479, 92.4%) were in HW equilibrium.

Most of the out-of-equilibrium variants observed in the Iberian Roma cohort are also present in MGP+1000G population with the same single alternative allele (5,068 variants, 87%). For these variants, a chi-square comparison test has been carried out to assess the differences in allele frequencies between the Iberian Roma cohort and the Spanish MGP+1000G populations. In this comparison, allelic frequencies distribution was significantly different (P < 0.05) for 3,601 variants (71.1%, Supplementary Table 1), while there were no differences in 1,267 variants (25%). The rest of the variants (200, 3.9%) could not be assessed for numerical restrictions (NA). Amongst the small percentage of variants that clearly deviated from equilibrium (P < 0.05), some are known pathogenic variants described in the general population (not being Roma private variants). This is the case for the *GJB2* p.Met34Thr variant, which causes autosomal non-syndromic deafness. Its genotypes distribution in the Iberian Roma population is also significantly different when compared with MGP+1000G population (P < 0.05). *HOGA1* p.Gly287Val, responsible for autosomal recessive hyperoxaluria and *HINT1* p.His112Asn), cause of autosomal recessive axonal neuropathy are also in disequilibrium in the Iberian Roma, not being present in the MGP+1000G populations.

### 6. Pathogenic alleles

Forty one individuals in the cohort (34%) were carriers of a known pathogenic Roma founder variant. For some of these variants, the carrier rate was as high 10.9 % for the *GJB2* p.Trp24Ter, being very rare in the general European population. The *GJB2* p.Trp24Ter is a frequent cause of autosomal recessive non syndromic deafness among the Roma[50], with an allele frequency in our cohort of 5.5%, higher than previously described in other Roma groups[51]. Three individuals are heterozygous carriers of two different founder pathogenic variants each. Table 4 shows the most frequent Roma founder variants found in the cohort. Moreover, some pathogenic variants, described in the general population, not private to the Roma, have a remarkably high allele frequency in our cohort. Hundred and sixty variants classified as pathogenic or likely pathogenic by ClinVar (to date September 2022) were present in our study population (Supplementary Table 2). These data should be considered cautiously, since some of the variants classified as pathogenic or likely pathogenic by ClinVar correspond risk factors or susceptibility alleles rather than disease causing variants. Moreover, assertion criteria are not always provided for the listed variants. Of those 160 variants, 152 were more frequent in our cohort than in the general population of gnomAD[4]. A total of 113 individuals (95% of the cohort) carry two variants or more classified as pathogenic or likely pathogenic by ClinVar and 18 individuals (15% of the cohort) were carriers of seven or more pathogenic or likely pathogenic variants each. Three individuals are carriers of the *SPG7* p.Leu78Ter variant, known to cause autosomal recessive hereditary spastic paraplegia[52], with an allele frequency of 1.5% in our cohort, much higher than the 0.2% and 0.01% allele frequencies in the South Asian and European subpopulations of Gnomad v3.1.2[4, 53] respectively. In fact, this variant has previously been reported in a Spanish Roma family[54].

**Table 4.** Most frequent Roma founder pathogenic variants identified in the cohort. For each variant, the following information is provided, **Gene**: HGNC symbol of variant carrier gene; **Change**: Change at DNA or RNA level according to HGVS nomenclature; **Genotypes**: R/R refers to homozygous reference genotype counts in the Iberian Roma Cohort, R/A refers to heterozygous genotype counts in the Iberian Roma Cohort, A/A refers to homozygous alternative genotype counts in the Iberian Roma Cohort; **Carrier rate**: percentage of individuals in the Iberian Roma Cohort who carry the variant; **Frequency**: *Freq R* refers to the reference allele frequency in the Iberian Roma Cohort, *Freq A* refers to the alternative allele frequency in the Iberian Roma Cohort and *MAF* refers to the Minor Allele Frequency in the Iberian Roma Cohort; **1000G**: *All* refers to the alternative allele frequency in the whole population of 1000 genomes Project Database (Phase 3) and *Eur* refers to the alternative allele frequency in the European Population of 1000 genomes Project Database (Phase 3); **Exac**: *All* refers to the alternative allele frequency in the whole population of Exome Aggregation Consortium (ExAC); **Associated Disease**: variant-related disease; **Reference**: bibliographic reference describing the association between the variant and the disease; **rs**: Reference single nucleotide polymorphism ID in dbSNP (https://www.ncbi.nlm.nih.gov/snp/); **Variant**: variant in format *chromosome : position within the chromosome* (human genome build Grch37; hg19) *: Reference Allele : Alternative Allele*; **Clinical Significance**: clinical significance according to ClinVar (https://www.ncbi.nlm.nih.gov/clinvar/)

| Gene | Change | Genotypes |  |  | Carrier rate (%) | Frequency |  |  | 1000G |  | Exac | Associated disease | Reference | rs | Variant | Clinical Significance |
| --- | --- | --- | --- | --- | --- | --- | --- | --- | --- | --- | --- | --- | --- | --- | --- | --- |
|  |  | R/R | R/A | A/A |  | Freq R | Freq A | MAF | All | Eur |  |  |  |  |  |  |
| GJB2 | p.Trp24Ter | 106 | 13 | 0 | 10.924 | 0.945 | 0.055 | 0.055 | 0.0004 | 0 | 0.001 | Deafness | Schrauwen, 2019 | rs104894396 | 13:20763650:C:T | Pathogenic |
| SLC12A3 | c.1180+1G>T | 110 | 8 | 1 | 7.563 | 0.958 | 0.042 | 0.042 | - | - | 0 | Gitelman syndrome | Coto, 2004 | rs749098014 | 16:56912074:G:T | Pathogenic |
| EIF2AK4 | p.Pro1115Leu | 112 | 7 | 0 | 5.882 | 0.971 | 0.029 | 0.029 | - | - | 0 | Pulmonary venoocclusive disease 2 | Navas-Tejedor, 2017 | rs774906916 | 15:40295502:C:T | Pathogenic |
| BIN1 | p.Arg234Cys | 116 | 3 | 0 | 2.521 | 0.987 | 0.013 | 0.013 | - | - | 0 | Centronuclear myopathy 2 | Cabrera-Serrano, 2018 | rs777176261 | 2:127821221:G:A | Likely pathogenic |
| SPG7 | p.Leu78ter | 116 | 3 | 0 | 2.521 | 0.987 | 0.013 | 0.013 | - | - | 0 | Hereditary spastic paraplegia | Sanchez-Ferrero, 2013 | rs121918358 | 16:89576947:T:A | Pathogenic |
| GALK1 | p. Pro28Thr | 117 | 2 | 0 | 1.681 | 0.992 | 0.008 | 0.008 | - | - | 0 | Galactokinase deficiency with cataracts | Kalaydjieva, 1999 | rs104894572 | 17:73761136:G:T | Pathogenic |
| BCKDHA | p.Trp391Gly | 117 | 2 | 0 | 1.681 | 0.992 | 0.008 | 0.008 | - | - | 0 | Mapple syrup urine disease | Quental, 2008 | rs398123489 | 19:41916544:C:- | Pathogenic |
| CD8A | p.Gly111Ser | 117 | 2 | 0 | 1.681 | 0.992 | 0.008 | 0.008 | - | - | 0 | CD8<sup>+</sup> T-cell immunodeficiency | Mancebo, 2008 | rs121918660 | 2:87017523:C:T | Pathogenic |
| ITGA8 | c.2982+2T>C | 117 | 2 | 0 | 1.681 | 0.992 | 0.008 | 0.008 | - | - | - | Bilateral renal agenesis | Humbert, 2014 | rs587777279 | 10:15573047:A:G | Pathogenic/Likely pathogenic |
| FANCA | p.Gln99ter | 117 | 2 | 0 | 1.681 | 0.992 | 0.008 | 0.008 | - | - | - | Fanconi anemia | Callén, 2005 | rs1057516430 | 16:89877468:G:A | Pathogenic |
| SGCG | p.Cys283Tyr | 118 | 1 | 0 | 0.840 | 0.996 | 0.004 | 0.004 | - | - | - | Muscular dystrophy, limb-girdle, autosomal recessive 5 | Piccolo, 1996 | rs104894422 | 13:23898652:G:A | Pathogenic/Likely pathogenic |
| CHRNE | p.Glu443LysfsTer64 | 118 | 1 | 0 | 0.840 | 0.996 | 0.004 | 0.004 | - | - | 0 | Myasthenic syndrome, congenital, 4C, | Kalaydjieva, 2001 | rs763258280 | 17:4802186:C:- | Pathogenic |
| LTBP2 | p.Arg299Ter | 118 | 1 | 0 | 0.840 | 0.996 | 0.004 | 0.004 | - | - | 0 | Weill-Marchesani syndrome 3/ Glaucoma | Morlini, 2018 | rs121918355 | 14:75022332:G:A | Pathogenic |
| MANBA | c.2158-2A>G | 118 | 1 | 0 | 0.840 | 0.996 | 0.004 | 0.004 | - | - | 0 | Mannosidosis, beta | Brozkova, 2020 | rs772852668 | 4:103556204:T:C | Pathogenic |
| SHOX | p.Ala170Pro | 118 | 1 | 0 | 0.840 | 0.996 | 0.004 | 0.004 | - | - | - | Léri-Weill dyschondrosteosis/Langer mesomelic dysplasia | Barca-Tierno, 2011 | rs397514461 | X:601577:G:C | Pathogenic |
| ACADS | p.Glu104del | 118 | 1 | 0 | 0.840 | 0.996 | 0.004 | 0.004 | - | - | 0 | Short chain acyl-CoA dehydrogenase deficiency | Lisyová, 2018 | rs387906308 | 12:121174884:GGA:- | Pathogenic/Likely pathogenic |
| SH3TC2 | p.Arg1109ter | 118 | 1 | 0 | 0.840 | 0.996 | 0.004 | 0.004 | - | - | 0 | Charcot Mary Tooth 4C | Gooding, 2005 | rs80338934 | 5:148389835:G:A | Pathogenic |
| GLB1 | p.Arg59Hys | 118 | 1 | 0 | 0.840 | 0.996 | 0.004 | 0.004 | - | - | 0 | GM1 gangliosidosis | Santamaria, 2007 | rs72555392 | 3:33114105:C:T | Pathogenic/Likely pathogenic |

### 7. Structural variants and adaptive mechanisms

Structural variants (SV) in the population are thought to have a role in adaptability and response to external pressures, with a great potential to lead to rapid evolution[55]. They are possibly involved in phenotypic diversity and disease susceptibility. We analysed the genomic regions harbouring SV that are specific to the Iberian Roma, and the genes present in these regions. Our analysis shows significant enrichment of deletions, inversions, insertions and duplications, but not translocations (Supplementary Table 3). Gene Ontology terms such as those related to inflammation, immune response and defence, including antigen presentation and processing, and cell-cell adhesion are significantly enriched (FDR adjusted p-value < 0.05). Keratinization and epidermal development were also significantly enriched in SV in our study population (FDR adjusted p-value < 0.05). Of note, 68% of the individuals in the cohort had SV involving KRTAP1-3, KRTAP5-7 or KRTAP5-8, encoding for keratin associated proteins. When we look specifically at inversions, over a 100-fold enrichment is observed in gene clusters related to oxygen and gas transport, and in particular to haemoglobin and haptoglobin complexes formation (Supplementary Table 3).

## DISCUSSION

The Roma constitutes an ethnic group with a unique demographic history and cultural tradition that that distinguish them from neighbouring populations. Nowadays, we are witnessing an exponential growth of the diagnostic and therapeutic options for genetic disease, which makes it even more necessary to provide this population group with the resources to take advantage of the recent developments. In this work we have developed a database of population variant frequency in the Iberian Roma to help in the variant assessment processes during genetic diagnosis.

We have relied on the self-definition of the individuals as Roma for their inclusion in the study. Our PCA analysis shows a perfect clustering of the samples together and apart from other non-Roma populations, in continuity with the Spanish and European clusters, supporting the accuracy of the classification of individual ethnicity based on self-identity (Figure 2). It is interesting to note these clusters do not overlap after five centuries of coexistence, although we know from previous studies that there has been admixture throughout the years[46]. However, there is a clear overlap with other non-Iberian Roma populations such as the North/Western Roma. The high number of RoH identified can be partially explained by the demographic history of the Roma with successive bottlenecks and reduced effective population size, while the presence of significantly long RoHs throughout the genome confirms the persistence of current and recent inbreeding in the population. These data are in keeping with the results of recent anthropological studies that showed consanguineous marriages amongst the Spanish Roma are decreasing in recent years, although still frequent[56].

Previous population studies have shown great numbers of SV, involving 4.8–9.5% of the genome, having around 100 genes completely deleted without apparent phenotypic consequences[57]. SV are thought to be related to adaptive evolution responding to environmental pressures and contributing to human diversity but also to disease susceptibility[58]. Our analysis of SV suggests the Iberian Roma have developed adaptive mechanisms involving immune response, similarly to other populations[55, 59]. More striking is the finding of significant SV enrichment in gene clusters related to keratinization and epidermal growth, suggesting the development of adaptive mechanisms related to climate and environmental stressors. Keratinization is associated with the evolution of hair in mammals. This gene family has evolved under selection among mammals as a response to environmental pressures to hair structure[60]. Interestingly, similar findings have been reported in Indian subpopulations[61]. Enrichment in SV involving other keratin related genes (KRTAP9-2, KRTAP9-3, and KRTAP9-8) have been described to be specific to African populations[55]. While these results show strong statistical significance, we must bear in mind that exome sequencing carries significant bias for the analysis of SV, missing most of the non-coding regions.

We have observed that some variants, including known pathogenic variants, are more common in the Iberian Roma than would be expected according to Hardy-Weinberg equilibrium. This deviation could be explained by small population size as well as genetic drift and selective coupling. Knowledge of the most prevalent pathogenic variants in a population facilitates diagnosis, more so in the field of rare disease, where phenotypes overlap and the process of finding a molecular diagnosis is often complex. For the Roma, this information is particularly helpful, as it is often one or a few variants that causes the majority of the cases of a disease in the population. However, in this study, we have seen a high carrier rate for some non-Roma pathogenic variants that were not known to be prevalent among the Roma. In fact, some of these variants have a much higher carrier rate among the Iberian Roma than in the general European population. Admixture and gene flow can explain the transmission of a specific allele from a host population (p.e. general European) to Iberian Roma. Genetic drift and selective coupling are possible mechanisms causing an increase of the new allele in the Iberian Roma, however, ethnicity is often not registered in clinical notes, therefore, it is possible that the original descriptions of some of these pathogenic variants, where the ethnic background of patients is not mentioned, correspond to Roma patients. The relatively small sample size analysed may not reflect accurately the actual prevalence of these conditions in the population, however, for many of them, figures shown are not too different from those seen in other Roma subpopulations.

The absence of a cohort of Roma ancestry in population databases of genetic variant frequencies often hinders a precise diagnosis for patients of this ethnicity. The open access database generated in the course of this study will hopefully contribute to improve genetic diagnosis in the Roma by increasing the resources available to this population.

## METHODS

### Recruitment of participants and sample collection

Inclusion criteria were being over the age of 18, self-reported Roma ethnicity of both parents and four grandparents and understanding and signing informed consent. Exclusion criteria were having a first degree relative already included in the study and having a known hereditary monogenic disease, a neurodegenerative condition of unknown cause or other disorder estimated by the researchers to be likely genetic. Vascular risk factors such as diabetes or hypertension were not considered a cause for exclusion from the study. Minimal demographic information was gathered from each volunteer including place of birth of the individual, place of birth of both parents, family history of disease and age at sample collection. A blood sample was taken from each volunteer between 2017 and 2020, for DNA extraction. Additionally, archive DNA samples from healthy Roma individuals were provided by Biobank Galicia Sur Health Research Institute (PT13/0010/0022) among other collaborations in Spain. For these samples, no demographic or biographic information was available. Samples were de-identified for sequencing and information was pseudo-anonymized and stored in a secure physical drive with restricted access under the custody of the researchers. IRB approval was obtained from bioethics and scientific committees of *Hospitales Virgen del Rocio-Macarena -Junta de Andalucía, Consejería de salud, igualdad y políticas sociales. (V°B° CEI33160037)*

### QC and exome sequencing

All DNA samples went through quality control before sequencing. Samples were run in a 1% agarose gel to ensure integrity. For purity control, absorbance was checked using Qbit, discarding samples with a A260/A280 below 1.7 or above 2. Two different capture kits were used for exome sequencing; MedExome capture was used for the first 20 samples, spanning a ∼47Mb target region. For the latter 99 samples, Xgen-exome-research-panel v1.0 was used, spanning a ∼39Mb target region. A Nextseq 500 Illumina platform was used for sequencing.

### Sequencing Data analysis

Two pipelines for processing the raw sequences (FastQ files) were developed. One for the discovery of SNVs and small indels (<50bp) and another for the discovery of structural variants (>=50bp).

SNV and small indel pipeline is based on GATK best practices[62] for a cohort study. For each sample, *FASTQC*[63] was used to assess quality of raw data and *fastp*[64] was run for quality pre-processing so that clean data is provided to downstream analysis. Then, filtered sequence reads were aligned to the reference human genome build hs37d5 (hg19) by using the BWA alignment tool[64]. The obtained mapped reads (BAM files) are then sorted by *samtools*[65] and duplicate reads are marked to mitigate biases introduced by data generation steps such as PCR amplification by means of *Picard tools* (“Picard Tools - By Broad Institute”). BAM files are later analysed in terms of QC using in-house scripts and the ngsCAT tool[66]. Then, *Base Quality Score Recalibration* (BQSR) is applied in a two-step procedure through GATK *BaseRecalibrator* and *ApplyBQSR* tools[67] with the aim of detecting and correcting for patterns of systematic errors in the base quality scores. After BQSR, per-sample variants (SNVs and small Indels) were identified through GATK’s *HaplotypeCaller* tool, by generating a GVCF file per sample. Then, a joint genotyping taking into account all samples in the cohort is run through GATK’s *GenomicsDBImport* and *GenotypeGVCF* tools. This way, the different records are merged together in a sophisticated manner, obtaining a set of joint-called SNPs and indels in a single multisample VCF file. With the aim of reducing putative false positives, a variant filtering step is run through the GATK’s Variant Quality Score Recalibration (VQSR) procedure restricted to the union of the two exome captures, producing a set of high-quality variants. This dataset is then annotated using the Cellbase database[43]. Finally, the set of high-quality variants is transformed into a set of counts of variants for the whole cohort which are inserted in the IRPVS database.

Structural variants (>=50bp)[68] pipeline is based on GRIDSS v2.7.3 software[69]. In order to characterize Iberian Roma-specific SVs, a Panel of Normal (PoN) SVs predicted using the same protocol in 138 samples belonging to the Navarra 1000 Genomes Project NAGEN1000 (“Proyecto Genoma Navarra NAGEN 1000 Navarra”) (https://www.navarrabiomed.es/en/research/projects/nagen1000) was used. This way, we excluded from our Iberian Roma cohort the SVs found in three or more samples belonging to the PoN, according to GRIDSS recommendations. Finally, using Cellbase database, the list of genes affected by the remaining set of SVs was extracted . For deletions and duplications, we considered genes falling between the boundaries of the SVs. In the case of inversions, insertions and translocations, we considered only genes falling in the SV breakpoints. Finally, we performed a gene ontology[70] functional enrichment analysis with PANTHER[71] over the set of genes affected by SVs in 10 or more samples in our cohort to get enough representation of SVs in the Iberian Roma population

### Comparison with other populations

For the analysis of Roma Iberian population in terms of overlapping with other populations for SNVs and small indels, three main reference datasets have been used: the 1000 Genomes Project[72], the Spanish population dataset from the Medical Genome Project[3] and the non-Iberian Roma dataset[46]. For the analysis in terms of autozygosity, the 1000 genomes and MGP datasets have been used. The Spanish population, sequenced in the context of the Medical Genome Project (http://www.clinbioinfosspa.es/content/medical-genome-project) includes 267 healthy, unrelated exome samples of Spanish origin (EGA, accession: EGAS00001000938) in a multisample VCF file[3]. We refer to this population as MGP.

For the 1000 genomes project, a total of five human macro-populations were used in this study, which included 661 African samples (YRI Yoruba in Ibadan from Nigeria, LWK Luhya in Webuye from Kenya, GWD Gambian in Western Divisions in the Gambia, MSL Mende from Sierra Leona, ESN Esan in Nigeria, ASW Americans of African Ancestry in SW USA, ACB African Caribbeans in Barbados), 347 Ad Mixed American samples (MXL Mexican Ancestry from Los Angeles USA, PUR Puerto Ricans from Puerto Rico, CLM Colombians from Medellin Colombia and PEL Peruvians from Lima Peru), 504 East Asian samples (CHB Han Chinese in Beijing China, JPT Japanese in Tokyo Japan, CHS Southern Han Chinese, CDX Chinese Dai in xishuangbanna China and KHV Kink in Ho Chi Minh City Vietnam), 489 South Asian samples (GIH Gujarati Indian from Houston Texas, PJL Punjabi from Lahore Pakistan, BEB Bengali from Bangladesh, STU Sri Lankan Tamil from the UK and ITU Indian Telugu from the UK) and 503 European samples (CEU residents of UTAH, TSI from Tuscany in Italy, FIN Finnish from Finland, GBR British from England and Scotland and the IBS from Spain). The genome sequences of all 2,504 individuals corresponding to the five super populations were downloaded from the 1000 genomes web page (https://www.internationalgenome.org/), last accessed July 31, 2019 in multisample variant calling format (VCF).

The non-Iberian Roma dataset includes 30 samples belonging to four main migrant groups: 10 Balkan, 5 Vlax, 10 Romungro and 5 North/Western Roma coming from four countries: Macedonia, Hungary, Lithuania and Ukraine. FASTQ files where downloaded from EGA database (accession EGAD00001006024) and processed similar to Iberian Roma cohort (see Sequencing Data analysis section) to obtain VCF files.

This way and using our Iberian Roma dataset, a total of 2,920 samples have been studied. When comparing populations and with the aim of reducing bias in number of variants found per individual due to heterogeneity of data (genomes and exomes with different captured regions), the analysis was restricted to the same regions of the 2,920 samples, namely, common captured regions in Iberian Roma and MGP populations and exonic regions from RefSeq (downloaded in October, 2019). As a consequence, studied regions cover ∼ 36.5 Mb.

### PCA generation

To compare genomic structure of the Iberian Roma population against the aforementioned populations, a Principal Component Analysis (PCA) was performed with PLINK v1.90. The first two dimensions were considered and drawn using R package ggplot2.

### Test for selection

In order to study possible deviations from the Hardy–Weinberg equilibrium we used the R package HWChisqStats[73], a function for the fast computation of chi-square statistics (or the corresponding p-values) for a large set of genetic variants (typically SNVs from sites with a single alternative allele). This test allows to evaluate all categories of variants: autosomal, X chromosomal and pseudo-autosomal regions (PAR1 and PAR2) of the X-chromosome variants, with their own particularities. Additionally, a chi-square test was used to assess the significance of differences in allele frequencies in the Iberian Roma population when compared with the MGP+1000G populations.

### Autozygosity study

The autozygosity of the Iberian Roma population was studied by detecting the runs of homozygosity (RoH) of each sample using *bcftools roh*[74]. The standard parameters recommended by the tool were applied to obtain those runs. The generated regions of autozygosity were then compared against homozygosity in 1000 genomes and MGP datasets, both in terms of their averaged length and the proportion of the autosomal genome (Froh). The Wilcoxon non-parametric statistical test was performed to determine whether homozygosity was significantly different among these datasets.

## DATA AVAILABILITY

Sequence data has been deposited at the European Genome-phenome Archive (EGA, see https://ega-archive.org/ last accessed December 5, 2022), under accession number EGAS00001006758.

## CODE AVAILAVILITY

IRPVS database code is available at https://github.com/babelomics/csvs/tree/gpvs. All the bioinformatics tools used are publicly available and referenced accordingly.

## ACKNOWLEDGEMENTS

We thank all the participants and particularly the community of “El Vacie” (Sevilla), for taking part in the study and for their necessary insight. We also thank David Comas (Universitat Pompeu Fabra, Barcelona) for his contribution, and Luba Kalaydjieva, Gianina Ravenscroft and Nigel Laing (University of Western Australia) for their fruitful suggestions and revision of the manuscript.

## CONTRIBUTIONS

M.C.S., F.M. and C.P. contributed to the study concept and design, F.M, A.V., M.L.A.V., M.S., S.S., P.F., A.M.N.N., A.C., Y.M., D.A., R.U, J.G.., P.M., C.P., and M.C.S. contributed to the recruitment of participants and acquisition of data, J.P.F., F.M.O., R.C. and D.L.L. performed the data analysis. G.R. developed the IRPVS website and the corresponding database. J.P.F. and M.C.S. drafted the manuscript, J.G. reviewed the manuscript and provided valuable feedback. C.P., J.D. and M.C.S. supervised the study and reviewed the manuscript for important intellectual content. All authors approved the submission of this manuscript.

---

**Supplementary Figure 1.**
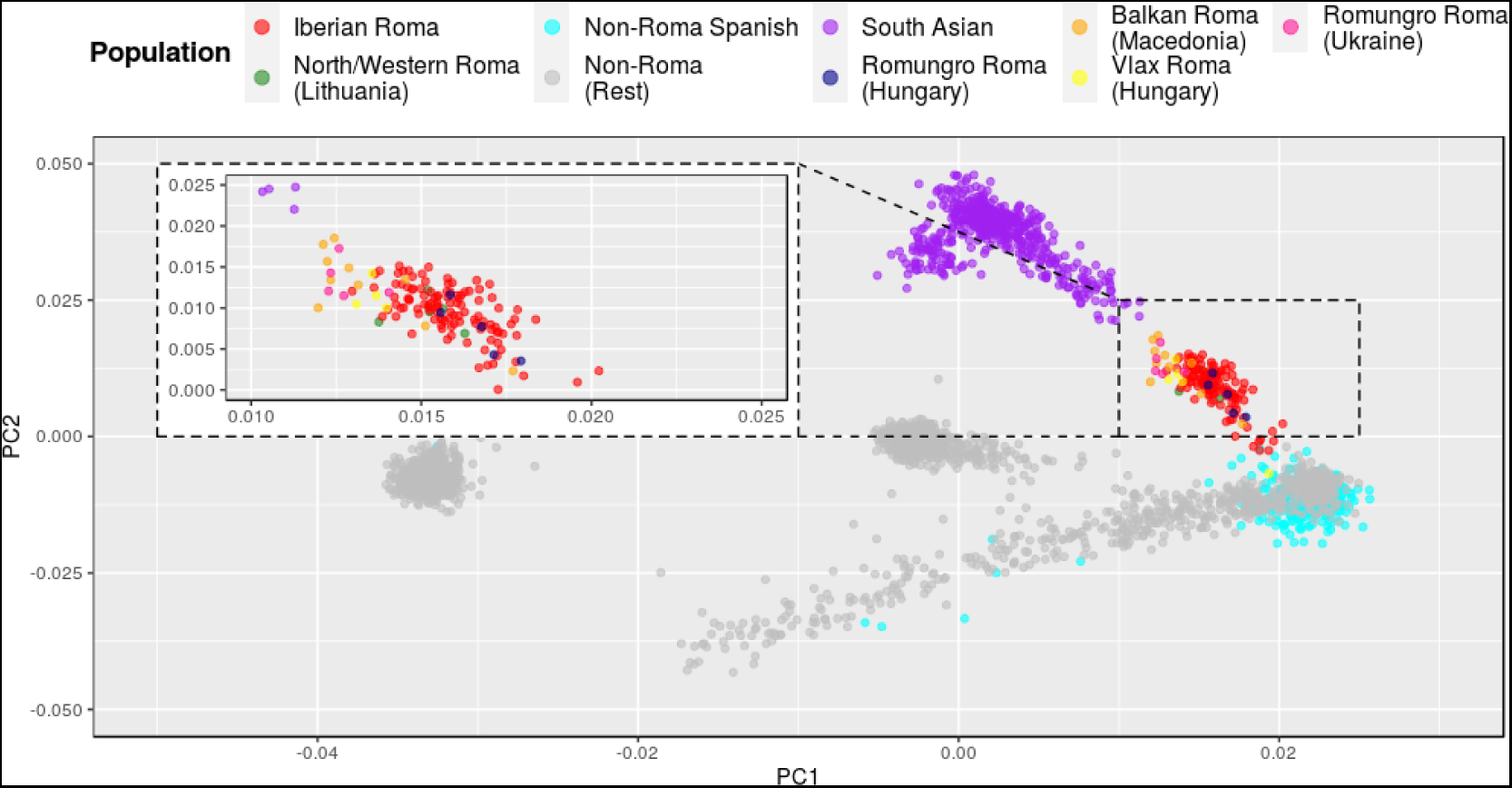
Principal Component Analysis (PCA) with a focus on Iberian and non-Iberian Roma population. PCA analysis performed using the 119 Iberian Roma samples included in this study as well as MGP, Thousand Genomes subpopulations and non-iberian European Roma restricted to common captured regions in Iberian Roma and MGP populations and exonic regions from RefSeq. The proximity and overlapping with the different migrant groups from non-iberian European Roma is detailed.

**Supplementary Figure 2.**
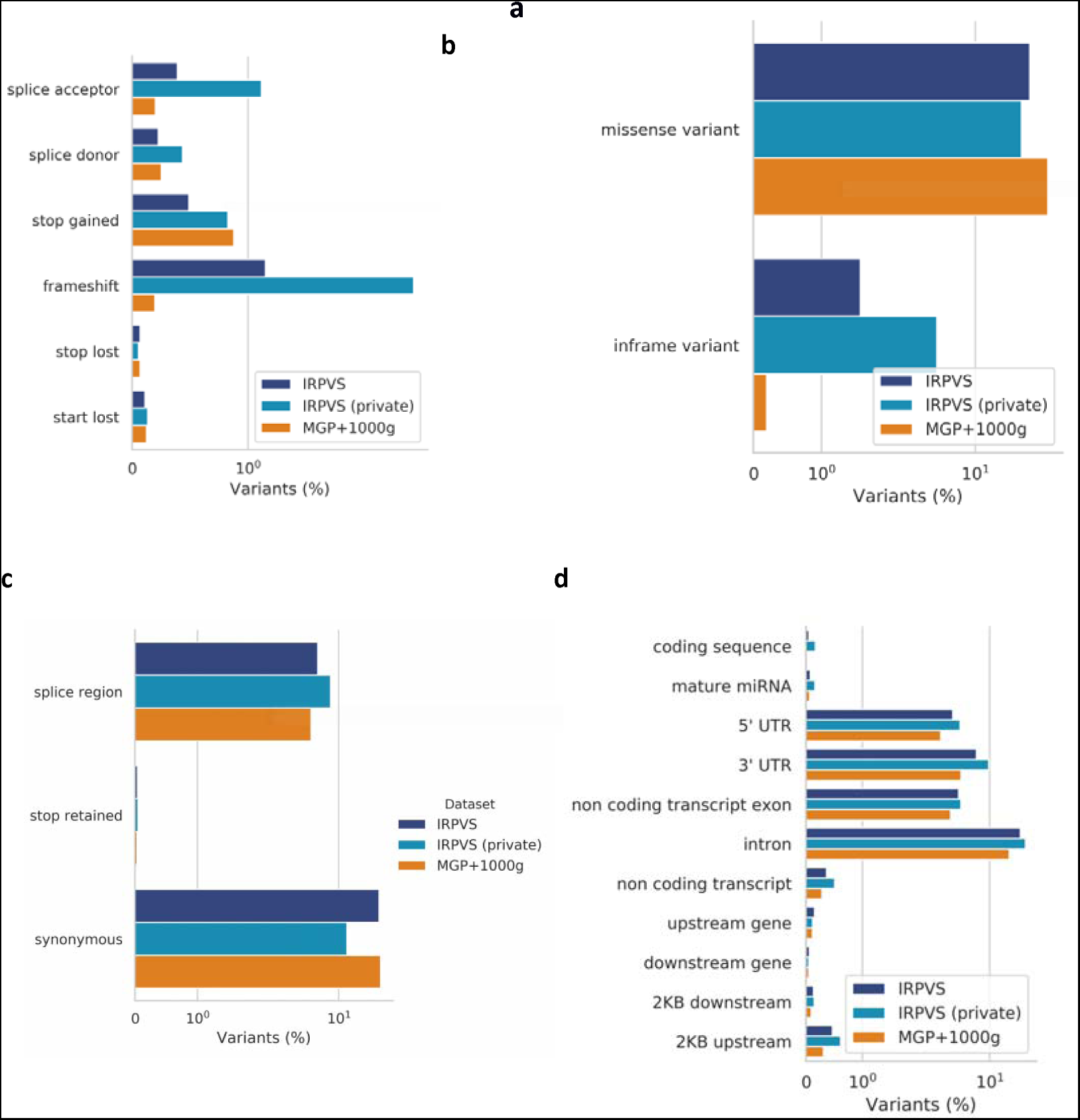
Distribution of variants per consequence types and impact. Distribution of variants in IRPVS (dark blue bar) and MGP+1000G (orange bar) according to Ensemble’s worst consequence type (the worst effect that the variant has on the set of transcripts) obtained through *Cellbase* and restricted to the union of the two exome captures of IRPVS data. Iberian Roma variants not present in MGP+1000G population (light blue bar) is also shown. Consequence type details are shown for each of the main categories: **a** HIGH, **b** MODERATE, **c** LOW and **d** MODIFIER. For visualization purposes, X-axes are in log scale and only terms with at least 0.03% of variants are shown. Details on consequence types (Sequence Ontology, SO, terms) are available at https://m.ensembl.org/info/genome/variation/prediction/predicted_data.html

**Supplementary Table 1. List of genomic variants in the study population which significantly (p < 0.05) deviate from Hardy-Weinberg (HW) equilibrium.** Table provided as excel file: Supplementary table 1.xlsx

**Supplementary Table 2. Pathogenic alleles by Clinvar present in the study population.** Table provided as excel file: Supplementary table 2.xlsx

**Supplementary Table 3. Gene Ontology terms more significantly involved by structural variants.** Table provided as excel file: Supplementary table 3.xlsx

